# Evaluation of linkage between health, education and social care administrative data for 21 million children in England

**DOI:** 10.64898/2026.09.16.26363127

**Authors:** Vincent Nguyen, Joseph Lam, Tony Stone, Ruth Blackburn, Ruth Gilbert, Milagros Ruiz Nishiki, Katie Harron

## Abstract

**Background:** Population-level administrative data cohorts hold huge potential for generating evidence to improve child health. Understanding the quality of these data is critical to enabling us to fully realise this potential. We aimed to evaluate linkage between children’s health, education and social care administrative records (ECHILD) to identify possible sources of bias.

**Methods:** We created four cohorts to understand the drivers of inclusion/exclusion within ECHILD. For each cohort, we calculated the linkage rate by counting the total number of eligible individuals who appeared in the ECHILD linkage spine and any of the corresponding hospital/school component datasets. We then assessed whether linkage rates varied according to sociodemographic characteristics (year of birth, sex, ethnicity, region).

**Results:** ECHILD currently captures 34.2 million patients and 25.3 million pupils born between September 1984 and March 2023, aged 0-38 years. Over 90% of children with hospital birth records linked to a school record (rising to 93% for those born from 2014 onwards); 88% of children attending school in 2001/02 linked to a patient record (rising to 96% for those born in 2019/20), resulting in a total of 21 million linked individuals. Substantial variation existed between sociodemographic groups, with those living in London, residing in more deprived areas, or with recorded ethnicity other than White being less likely to be included.

**Conclusions:** Population-level datasets such as ECHILD often under-represent specific groups. Irrespective of the mechanisms by which individuals are excluded (e.g. opt outs, linkage errors or not interacting with services), those in minoritised ethnic groups and those living in more deprived areas are most affected. Linkage evaluations are critical for understanding who is and who is not included in analyses of these data so that researchers can account for potential sources of bias within analyses.

**Highlights:**

- We evaluated linkage quality and population coverage within ECHILD, a national, longitudinal, linked health, education and social care data resource.
- ECHILD contains linked health and education records on 21.3 million children born from 1984 onwards; linkage rates were >90% for hospital birth cohorts and >88% for school inception cohorts.
- Linkage rates varied by ethnicity, deprivation and geography, with lower representation of minoritised groups and children living in London.
- Routine linkage evaluations are essential for quantifying selection bias and supporting valid use of population-level administrative data; our findings demonstrate the importance of transparent reporting of linkage quality for population data science research.

## Introduction

The UK has seen a serious decline in child health over recent decades. Infant mortality has ranked 28 out of 35 OECD countries, demand for child mental health services has become unsustainable, more than one in five children are overweight or obese by age five, and one in four have tooth decay.[1, 2] Childhood is a critical period in which the building blocks for lifelong physical and mental health are laid down, and the importance of the early years has been recognised in the UK government’s Opportunity mission, which aims to give all children the best start in life.[3] There is clear evidence that such investment will be cost-effective in enabling future adults to live long and productive lives.[4]

High-quality data on child health is key to achieving this goal, by providing evidence on the health needs and experiences of children and families with which to inform development of interventions and targeting of support. There is also increasing recognition of the need to move beyond traditional disciplinary boundaries, to understand social determinants of health, and the relationship between health and other areas of our lives, including households, schools, and the environments in which we live. For example, the Children’s Commissioner’s Office advocates for the linkage of health, education and social care data, citing the overwhelming body of evidence demonstrating that such linkage would be beneficial not only to individuals but to wider society through improved service delivery.[5]

A data resource that aims to meet these needs is ECHILD (Education and Child Health Insights from Linked Data). ECHILD brings together administrative data from health, education and social care for children in England born since September 1984 and their mothers.[6–8] It provides near population coverage, with detailed, linked, longitudinal information on ∼21 million individuals in England followed up for >350 million person-years. Linkage between children and their biological mothers enables insights into intergenerational factors, and facilitates sibling-control analyses to strengthen causal conclusions.

Whilst ECHILD is uniquely placed to help transform our understanding of child health, the challenges of using such administrative data for research are well understood: researchers need to carefully consider data quality, and specifically how to handle missing data, unmeasured confounding, non-representativeness, and other biases.[9] For example, we have previously demonstrated that based on a sample of ECHILD, linkage rates vary according to indices of deprivation and ethnicity, meaning that the data are less representative of particular groups of individuals within the population.[10]

A further challenge is that the source datasets capturing information from NHS and education services (primarily Hospital Episode Statistics [HES] and the National Pupil Database [NPD]) do not include the entire population. For example, the NPD only captures individuals attending state schools in England (plus those with Key Stage 4 or 5 results submitted by independent providers). Approximately 7% of children each year are in independent (private) schools and in 2024/25, an estimated 126,000 children are electively home schooled.[11, 12] Some children residing in England may attend schools in Scotland and Wales and therefore would not be captured in the NPD. Similarly, HES only captures individuals registered for NHS-funded services; individuals only using private healthcare would therefore not be captured, nor migrants or refugees, and those facing communication, language or cultural barriers.[13, 14] Individuals without a valid birth date recorded are excluded from the HES data in ECHILD. Individuals can also opt out of their health data from being included in ECHILD via the National Data Opt Out, which is a service that allows individuals to prevent their pseudonymised health records being used for healthcare research and planning.[15]

We aimed to conduct a comprehensive evaluation of ECHILD, by describing linkage rates and determining which groups of the population are and are not captured within the data resource. We examined four different cohorts of children and young people born from 1984 onwards to determine the proportion of individuals captured in HES who had a link to NPD, for i) 0-19 year olds born 1984 to 2017 with any HES record, and ii) individuals with a birth record in HES between 2003 and 2017. We also aimed to determine the proportion of individuals captured in NPD who had a link to HES, for a) any individuals with a record in NPD between September 1995 and August 2022, and b) individuals enrolled in school in the academic year 2001/02.

## Methods

### Data Sources

In England, information on a child’s journey through education and social care is recorded in administrative records in the NPD held by the Department for Education.[16] The NPD includes information on children’s school attendance and attainment, and children’s social care data (CSC).[17, 18] Within the NPD, a unique and anonymised child-level identifier called the Anonymised Pupil Matching Reference (aPMR) can be used to link data across different years of data collection.

NHS England holds demographic information about all patients accessing services funded by the NHS in England within the Personal Demographic Service (PDS), as well as all NHS-funded hospital contacts (captured in HES), mental health services, community services, maternity services, and birth and death registrations (the latter collated by the Office for National Statistics; ONS).[19, 20] A pseudonymised, unique identifier (the Token Person ID; TPI) can be used to follow individuals within and across the health datasets.

For simplicity, we will refer to all the datasets supplied by NHS England under the umbrella of “HES” and all the datasets supplied by DfE (including NPD, ILR and CSC records) under “NPD”. ECHILD brings together data from all of these sources (**Appendix 1**).

### Linkage and Opt Outs

Linkage between PDS and NPD records was conducted by NHS England, who used deterministic linkage of name, date of birth, sex and postcode to create an ECHILD linkage spine which allows researchers to map between aPMR and TPI (**Appendix 2**).[21] Within the linkage, TPIs were assumed to be unique but aPMRs were not: each aPMR was therefore linked to at most one TPI, but a TPI may have linked to more than one aPMR. This means that whilst each pupil could link to only one patient record, a single patient record could link to multiple pupil records. Where a single aPMR matched multiple candidate TPIs during linkage, no link was retained in the released linkage spine (due to NHS E processes that prevent uncertain linkages from being released).

Following linkage, NHS England removed any health data (including TPIs in the linkage spine) for participants with a National Data Opt Out registered at the point at which the linkage was completed. Data for unlinked NPD records and unlinked HES records are retained within ECHILD. This means that DfE records for individuals who have a health data Opt Outs are still captured in ECHILD, but would not be linkable to health data.

### Eligibility for linkage

We created four cohorts to understand the different drivers of inclusion/exclusion within ECHILD (**Table 1**). The first two cohorts (HES to NPD) allow us to examine the factors associated with not linking to NPD (i.e., linkage error, or no record of attendance at state schools due to private schooling, death, or immigration). The second two cohorts (NPD to HES) allow us to examine factors associated with not linking to HES (i.e., linkage error, no NHS interactions, and Opt Outs).

**Table 1:**
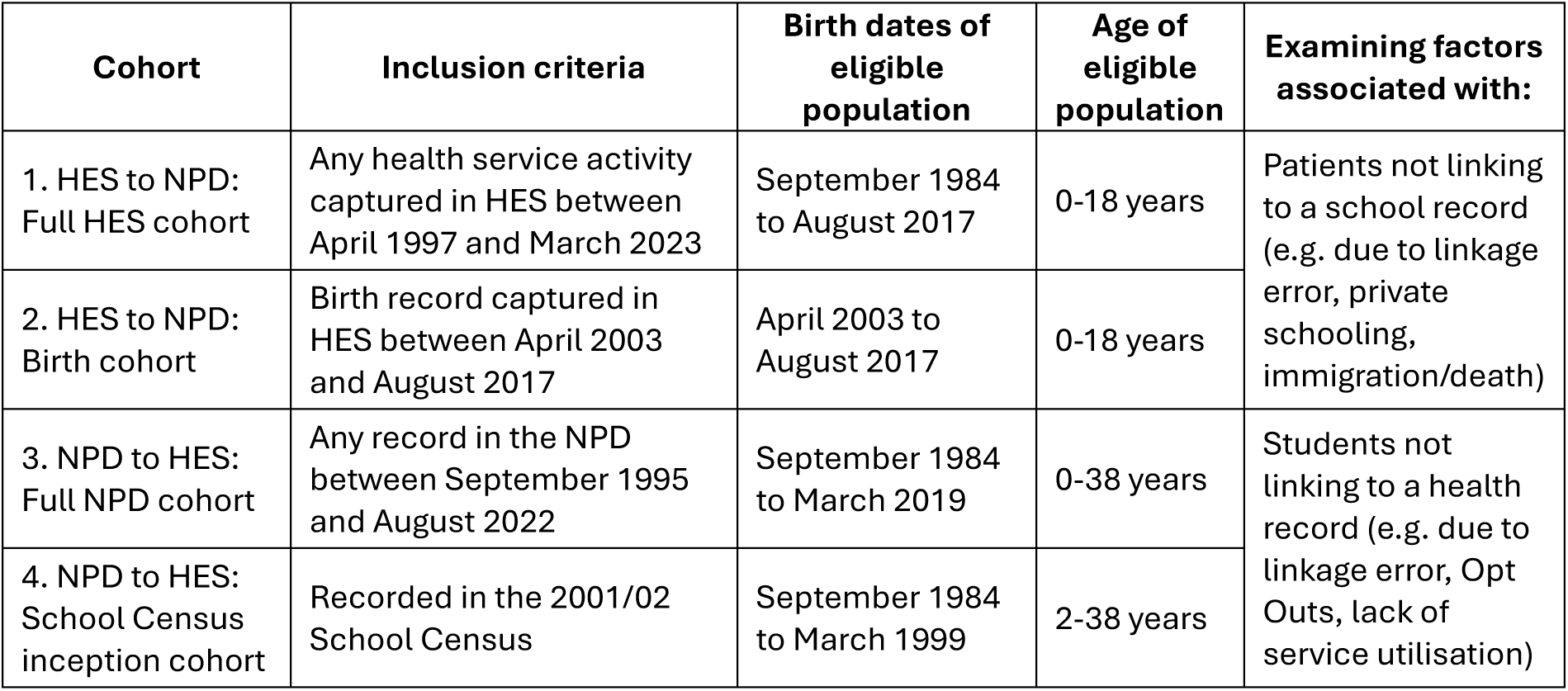
Definition of Cohorts used in analyses.

For each cohort, we restricted our denominator to those we expected to have been eligible for linkage (details for each cohort given below). Although ECHILD contains health data for mothers born prior to September 1984, these mothers would have been too old to link to the NPD; our evaluation therefore only included individuals born from September 1984 onwards. An overview of the cohorts is provided in **Figure 1**.

**Figure 1:**
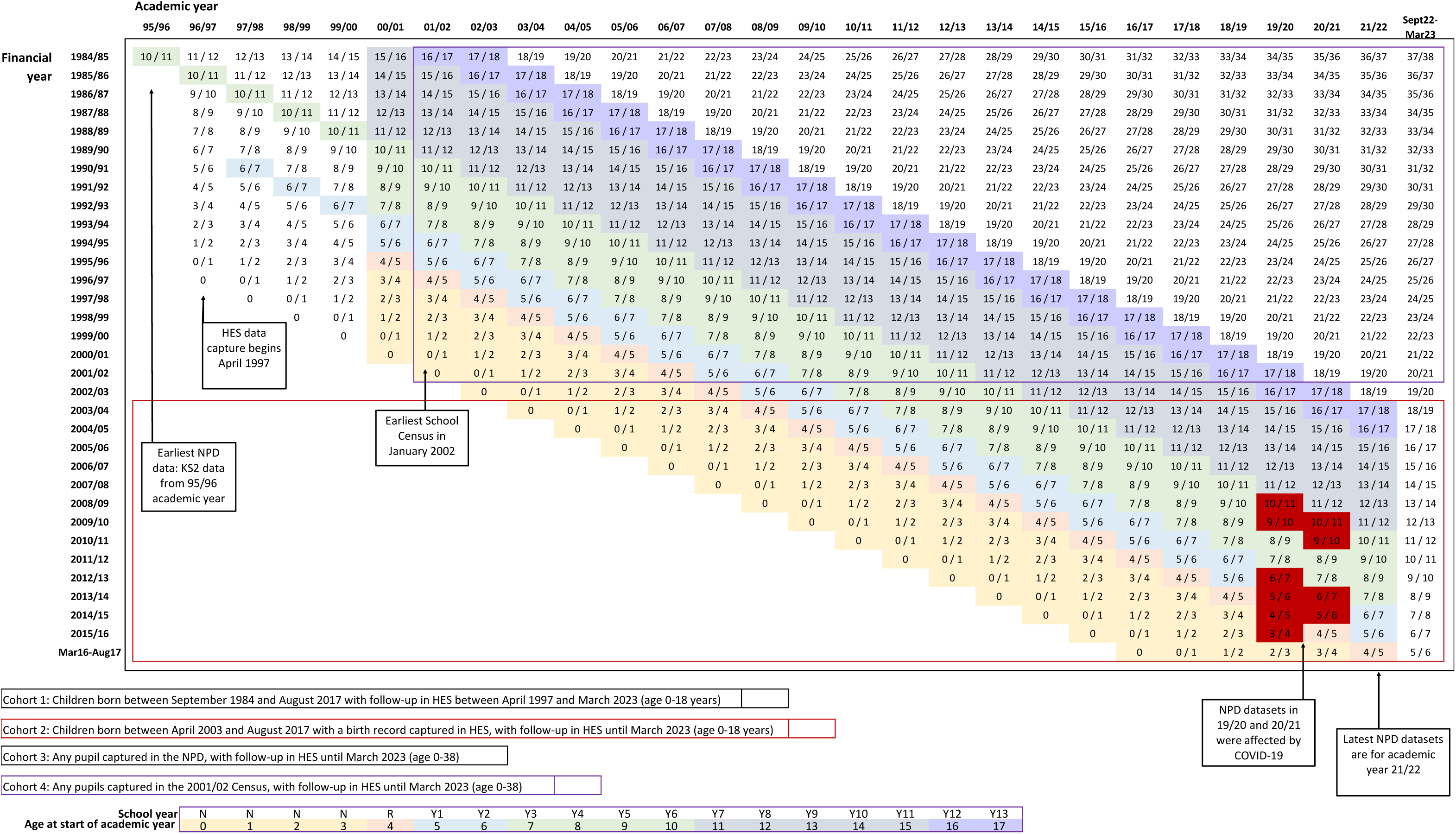
Overview of cohorts and follow-up in ECHILD. Numbers in boxes are age of individual during the academic year. Shading corresponds to school stages.

*Cohort 1*: HES to NPD linkage: Full HES cohort for children born between September 1984 and August 2017 with any health service activity captured in HES between April 1997 and March 2023 (aged 0-18 years).

In the NPD, the primary data source for school enrolments is the Census. The Spring Census is taken in January each year, and captures those enrolled at state nurseries, and primary, secondary and special schools. The first Census in NPD was for the academic year 2001/2002 (September to August), and the latest available Spring Census currently in ECHILD is for the academic year 2021/2022. The oldest cohort of children appearing in the census would have been entering Year 13 at age 17, i.e. born from September 1984 onwards. The youngest full cohort of children captured in the census would be those entering Reception (having turned 4 years of age) in 1^st^ September 2021 (i.e. born by the end of August 2017; though ECHILD also contains data for younger children attending government-funded childcare, it does not capture all children at these ages). To determine how many individuals captured in HES linked to an NPD record, we therefore restricted our HES cohort to those with birth dates between September 1984 and August 2017.

We further restricted this cohort to individuals with a first appearance in HES before the age of 19 (i.e. those who would have had a chance to be captured in NPD). Some children may have died prior to school age, but we did not systematically exclude deaths since the Early Years Census data will include some children from age 2 who were entitled to free childcare (and the numbers of deaths in children younger than 2 years of age is small). We were unable to account for those who migrated prior to starting school.

*Cohort 2:* HES to NPD linkage: Birth Cohort April 2003 to August 2017

Researchers using ECHILD often create birth cohorts, allowing them to follow individuals from birth, through childhood and into early adulthood.[22] Since October 2002, babies have been assigned NHS numbers at birth. From this time, linkage between birth records and subsequent patient records in HES is more reliable. We therefore further restricted our cohort to individuals with birth records within HES between April 2003 and August 2017.

Patients in Cohorts 1 and 2 should appear in the NPD unless they never attended a state school in England.

*Cohort 3:* NPD to HES linkage: Full school cohort for all pupils captured in NPD

To determine the proportion of pupils with a link to HES, we created a cohort of all individuals appearing in any NPD dataset (i.e., children with a valid aPMR).

*Cohort 4:* NPD to HES linkage: School inception cohort 2001/02

Analogous to birth cohorts, many researchers will create “school inception” cohorts in ECHILD, by restricting their population to individuals enrolled within school for a particular set of academic years and following them up longitudinally within HES.[23]

Since the earliest School Census was in the academic year 2001/2002, we created an NPD inception cohort by taking all those present in this academic year.

Pupils in cohorts 3 and 4 should appear in HES unless they had never had an interaction with NHS services, did not have a valid date of birth recorded in HES, or had a National Data Opt Out.

### Evaluation: linkage rates

For each cohort, we calculated the linkage rate by counting the total number of eligible individuals who appeared in the ECHILD linkage spine and any of the corresponding HES/NPD component datasets. We describe linkage between NPD and HES (rather than NPD and PDS), because ECHILD only holds attribute data for individuals within PDS who have a HES record; we would not have been able to apply eligibility criteria based on e.g. date of birth for PDS individuals without HES records.

For a subset of Cohort 2 (born between September 2003 and August 2004), we counted how many individuals appeared in the different NPD component datasets (School Census, KS4 / KS5 attainment, CiN, CLA, EYC, Alternative Provision, Individualised Learner Record [ILR]).

Finally, for a subset of NPD (those in the Spring Census 2021/22, from nursery to Year 13, born between 2003 and 2017), we additionally counted how many individuals linked back to their birth record in HES (either a birth registration record, a birth notification record, or an Admitted Patient Care record identified as being a birth episode).[8]

We use the term ‘linkage rates’ within this evaluation, but these percentages should not be interpreted as measures of linkage accuracy alone, as they represent inclusion more broadly within the linked data. This may be influenced by a combination of linkage accuracy, source dataset coverage, service utilisation, and National Data Opt Outs.

We assessed whether linkage rates varied according to sociodemographic characteristics (year of birth, sex, ethnicity, region), by comparing the percentage of individuals with these characteristics across linked and unlinked groups. Within HES, ethnicity is captured with 21 categories, and for the birth cohort we additionally described unlinked records according to birth characteristics (birthweight, gestational age and maternal age) and decile of the Index of Multiple Deprivation (IMD).[24]

In NPD, ethnicity is captured with 5 categories, and for the School Census cohort (Cohort 4), we additionally described unlinked records according to pupil characteristics (Free School Meals, Special Educational Needs and Disability, Mother Tongue [first language], National Curriculum Year Group and decile of Income Deprivation Affecting Children Index [IDACI]).

Characteristics were derived from the source dataset used to define cohort eligibility (HES for Cohorts 1–2; NPD for Cohorts 3–4). We did not prioritise one system’s demographics over the other; instead, analyses use the source-specific recording (e.g., HES-recorded ethnicity for HES cohorts; NPD-recorded ethnicity for NPD cohorts).

Where there were multiple conflicting values across records for the same individual within datasets, we used modal values.

### Evaluation: identifying potential false matches

To understand the potential for false links within ECHILD (where an individual in the NPD was linked to the wrong HES record), we enumerated the number of pupils with multiple links in the linkage spine (one TPI linked to multiple aPMRs) and described the characteristics of this group.

To quantify further potential false matches, we created a cohort of HES individuals we knew should not link to NPD (i.e. those in Cohort 2 who had died before school age, according to Civile Registrations of Death data) and described how many had a subsequent NPD record. This provides a minimal estimate of the false-match rate amongst the specific group of children who have died, but using these records as ‘negative controls’, allows us to explore the characteristics of those who are more likely to be falsely matched.

## Results

Within ECHILD, there were 34.2 million patients (distinct TPIs) and 25.3 million pupils (distinct aPMRs), for individuals born between September 1984 and March 2023, aged 0-38 years. NHS England reported that 5.8% of the aPMRs for which a link to a single TPI had been identified related to individuals with a registered Opt Out (but did not provide a demographic breakdown); these records were subsequently removed from the linkage spine. The resulting linkage spine contains 21.3 million unique pairs of records, representing 21.1 million distinct aPMRs linked to 21.3 million distinct TPIs (**Appendix 3**).

*Cohort 1*: HES to NPD linkage: Individuals born between September 1984 and August 2017 and captured in HES between April 1997 and March 2023 (aged 0-18 years).

Of the 22.9 million individuals within this cohort, 17.6 million (77%) linked to the NPD (**Figure 2**, **Appendix 4**). Lower linkage rates for those born 1997-2002 are likely explained by a lack of NHS number assigned at birth during these years. Lower linkage rates for those born in 1984 and 1985 may be explained by individuals moving to England on completion of schooling in other countries (i.e., appearing in HES for the first time between ages 16-18). Those with unknown ethnicity, no fixed abode or unknown geographies, and those living in London or in more deprived areas were less likely to link. Linkage rates were higher for females and those recorded as White British (**Figure 3**, **Appendix 5**).

**Figure 2:**
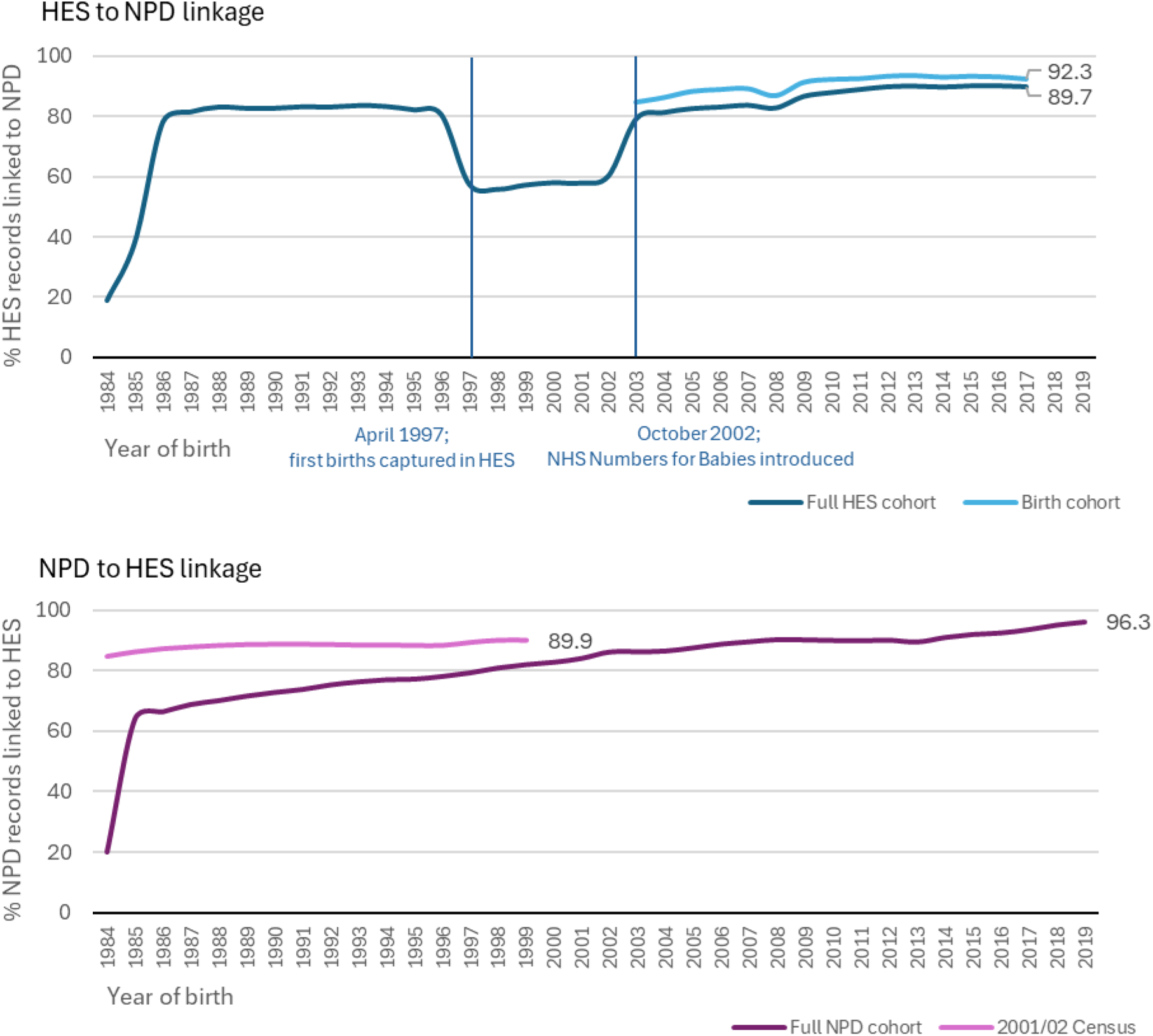
Top: Percentage of HES records (up to March 2023) linking to a pupil record (up to August 2022); Bottom: Percentage of NPD records linking to a patient record (bottom). Cohort 1: Full HES cohort for children born between September 1984 and August 2017 with any health service activity from April 1997 to March 2023. Cohort 2: HES birth cohort (individuals with a birth record available between April 2003 to August 2017). Cohort 3: Full NPD cohort (any pupils with an NPD record during academic years 1995/96 to 2021/22). Cohort 4: NPD School inception cohort (present in 2001/02 school census). Lower linkage rates in Cohort 1 for those born in 1997-2002 are likely explained by the fact that birth records were captured in HES for the first time in 1997 (when HES data collection began). Between 1997 and 2002, birth records and subsequent health records for the same child were more likely to have been assigned two different TPIs (prior to NHS numbers being assigned at birth from 2002 when NHS Numbers for Babies was introduced). In these cases, the linkage algorithm would only have allowed one link, or would not have returned a link where one NPD record linked to multiple TPIs. Hence, the low rate reflects a denominator inflated by duplicates and a numerator with records removed where duplicate TPI exist for the same person.

**Figure 3:**
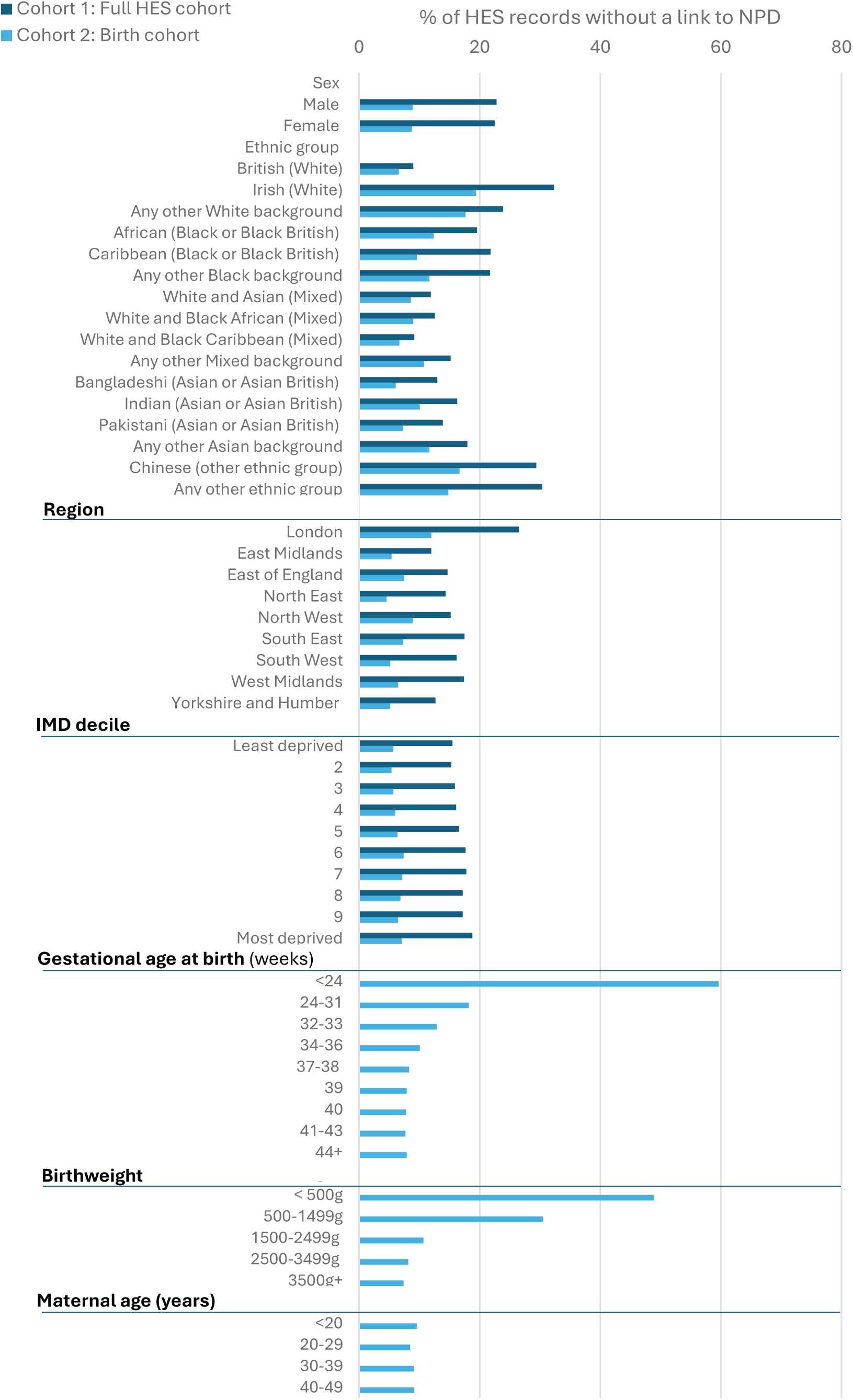
Characteristics of unlinked HES records from Cohort 1 (full cohort) and Cohort 2 (birth cohort)

*Cohort 2*: HES to NPD linkage: Birth Cohort April 2003 to August 2017 captured in HES between April 1997 and March 2023 (aged 0-18 years).

Of the 8.7 million individuals within the birth cohort, 90% linked to an NPD record (**Figure 2**, **Appendix 6**). Those with lower gestational ages and lower birth weight were less likely to link (**Figure 3**, **Appendix 5**). Of those born between September 2003-August 2004 (i.e. with 18 years of follow-up), the majority (83%) linked to a school Census record; 79% linked to a KS1 record, 81% linked to a KS4 record, 20% linked to CiN record, and 2% linked to a CLA record (**Table 2**).

**Table 2:** Percentage of Cohort 2 (born 2003-2017) linking to component NPD datasets up to academic year 2021/22 (age 0-19), by year of birth.

| Calendar year of birth | Max age at end of follow-up | N births | Census | Early Years Census | EYFSP | KS1 | KS2 | KS4 | KS5 | NCCIS | ILR | Absences | Exclusions | Alternative provision | Child in Need | Child Looked After |
| --- | --- | --- | --- | --- | --- | --- | --- | --- | --- | --- | --- | --- | --- | --- | --- | --- |
| *Age in years* |  |  | *2-19+* | *3/4* | *4/5* | *6/7* | *10/11* | *15/16* | *17/18* | *16/17* | *16+* | *all* | *all* | *all* | *all* | *all* |
| **2003** | 18 | 424,532<sup>1</sup> | 83 | 20 | 80 | 79 | 78 | 81 | 74 | 78 | 51 | 82 | 13 | 1 | 20 | 2 |
| **2004** | 17 | 570,785 | 84 | 48 | 81 | 80 | 79 | 82 | 73 | 79 | 48 | 83 | 13 | 1 | 21 | 2 |
| **2005** | 16 | 579,215 | 86 | 49 | 84 | 82 | 81 | 84 | 47<sup>5</sup> | 53<sup>5</sup> | 32<sup>5</sup> | 85 | 13 | 1 | 21 | 2 |
| **2006** | 15 | 596,118 | 87 | 50 | 84 | 83 | 82 | 55<sup>5</sup> |  |  |  | 86 | 13 | 1 | 21 | 2 |
| **2007** | 14 | 610,960 | 87 | 51 | 85 | 83 | 82 |  |  |  |  | 86 | 12 | 1 | 19 | 2 |
| **2008** | 13 | 642,111 | 85 | 50 | 83 | 81 | 53<sup>4</sup> |  |  |  |  | 84 | 9 | 1 | 18 | 1 |
| **2009** | 12 | 641,710 | 89 | 53 | 87 | 85 | 0<sup>4</sup> |  |  |  |  | 88 | 7 | 1 | 17 | 1 |
| **2010** | 11 | 659,158 | 90 | 55 | 88 | 86 | 29<sup>4</sup> |  |  |  |  | 89 | 4 | 1 | 16 | 1 |
| **2011** | 10 | 660,812 | 90 | 61 | 88 | 87 | 56<sup>5</sup> |  |  |  |  | 89 | 2 | 0 | 14 | 1 |
| **2012** | 9 | 662,700 | 90 | 65 | 89 | 58<sup>4</sup> |  |  |  |  |  | 89 | 2 | 0 | 13 | 1 |
| **2013** | 8 | 633,984 | 90 | 66 | 89 | 0<sup>4</sup> |  |  |  |  |  | 89 | 1 | 0 | 11 | 1 |
| **2014** | 7 | 629,777 | 90 | 67 | 59<sup>4</sup> | 30<sup>4</sup> |  |  |  |  |  | 87 | 1 | 0 | 10 | 1 |
| **2015** | 6 | 633,727 | 90 | 67 | 0<sup>4</sup> | 57<sup>5</sup> |  |  |  |  |  | 88 | 1 | 0 | 8 | 1 |
| **2016** | 5 | 633,577 | 90 | 67 | 30<sup>4</sup> |  |  |  |  |  |  | 59<sup>3</sup> | 0 | 0 | 6 | 1 |
| **2017** | 4 | 149,843<sup>2</sup> | 89 | 63 | 88 |  |  |  |  |  |  | 0<sup>3</sup> | 0 | 0 | 4 | c |
| **All** |  | **8,729,009** | **88** | **56** | **73** | **62** | **36** | **19** | **11** | **12** | **8** | **83** | **6** | **1** | **15** | **1** |

EYFSP: Early Years Foundation Stage Profile; KS: Key Stage; NCCIS: National Client Caseload Information System; ILR: Individualised Learner Record.
<sup>1</sup>Births from April-December; <sup>2</sup>Births from January-March; <sup>3</sup>Absence data not compulsory in reception year before 2024/25; <sup>4</sup>Birth years for which COVID-19 would partially/fully affected data collection during 2019/20 and 2020/21 academic years; <sup>5</sup>Only births from January-August would be expected to link.
“c” indicates suppression of small percentages.

*Cohort 3:* NPD to HES linkage: Any pupils captured in NPD

Of the 25.3 million individuals captured in the NPD at any time, 20.3 million (80%) linked to a HES record (**Figure 2**, **Appendix 7**). Linkage rates were lower for those born in 1984 - 1985 than later years; these pupils would only have been eligible for linkage from age 12 to 18, not all of whom would have used hospital services between these ages. Boys were more likely to link than girls; pupils with recorded Free School Meals or SEND provision were also more likely to link than those without (**Figure 4**, **Appendix 8**).

**Figure 4:**
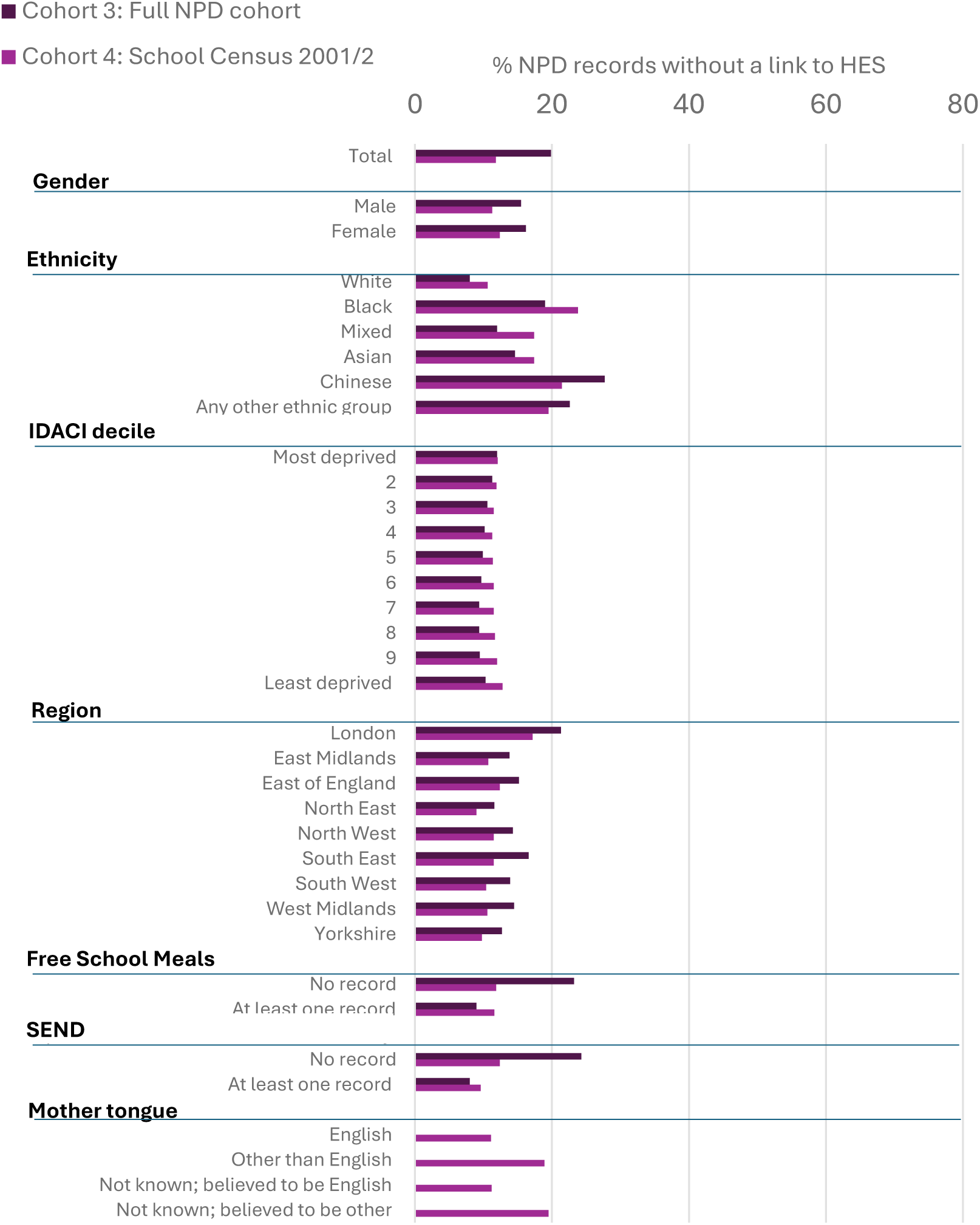
Characteristics of unlinked NPD records from Cohort 3 (full cohort) and Cohort 4 (census cohort)

*Cohort 4:* NPD to HES linkage: School inception 2001/02

Of the 7.6 million individuals captured in the NPD school inception cohort, 6.7 million (88%) linked to a HES record (**Appendix G**). Linkage rates were lower in those recorded with Black ethnicity, those living in London, and those whose first language (mother tongue) was other than English. Children with recorded SEND provision were more likely to link than those without (**Figure 5**, **Appendix 8**).

Finally, of the 8,181, 240 pupils present in the 2021/22 Census, 7,121,778 (87.1%) linked back to a birth record (83.0% to an APC birth record, 86.8% to a civil birth registration, and 78.9% to a birth notification).

### False matches

Of 21.0 million unique patients in the linkage spine, 124,533 (0.6%) linked to multiple aPMRs. The children with multiple aPMRs tended to be born from 2003 onwards, and be living in more deprived areas, London, or with no fixed abode, or be recorded in HES with African ethnicity (**Table 3**).

**Table 3:**
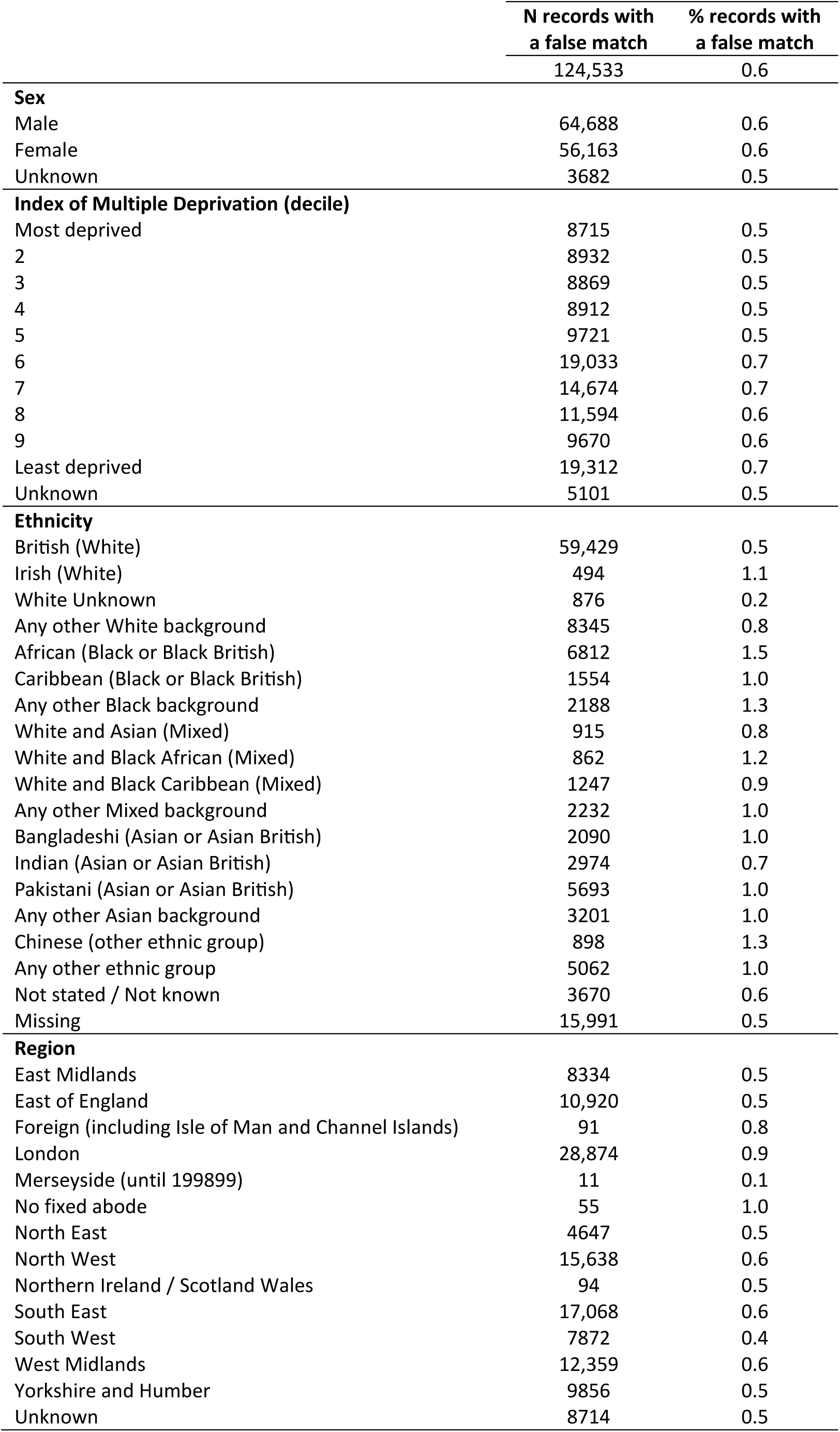
Characteristics of HES records that were linked to more than one pupil record (i.e., indicators of false-matches). Percentages are of the 21 million individuals in the linkage spine.

Of the 42,638 children in the birth cohort who died prior to school age, 70 (0.16%) linked to an NPD record after the date of death (and were therefore assumed to be false matches). These children tended to have missing data on key characteristics: sex (n=36), ethnicity (n=41), region (n=53), IMD (n=62), gestational age (n=43), birth weight (n=41), maternal age (n=43).

## Discussion

ECHILD represents a significant population data resource capturing information on health and education records for >34 million patients and >25 million pupils aged 0-38 years born from 1984. In our linkage evaluation, over 90% of children with birth records in HES linked to a school record (rising to 93% for those born from 2014 onwards), and 88% of children attending school in 2001/02 linked to a patient record (rising to 96% for those born in 2019/20), resulting in a total of 21 million linked individuals. We found that whilst it is possible to achieve high linkage rates, substantial variation exists between sociodemographic groups, with those living in London, residing in more deprived areas, or with recorded ethnicity other than White being less likely to be linked. These differences have implications for how national linked data can be used to generate evidence applicable to whole populations and specific subgroups.

While NHS England reported that 99.1% of submitted NPD records could be linked to a patient record, the inclusion rates observed within ECHILD were lower because the final dataset reflects not only linkage processes but also population coverage, service use, and National Data Opt Outs. The remaining 0.9% of unlinked records reflects a mixture of those with missing or poor quality identifiers, and individuals who were not captured in the PDS source dataset to start with.[25] The latter would include those never registered for or receiving NHS-funded services in England (including, for example, those attending school in England but residing in Wales or Scotland).

After National Data Opt Outs were applied to ECHILD, we observed linkage rates that were lower than the 99.1% reported by NHS England. The lower linkage rates we observed will in part be explained by datasets for which identifiers were not sent for linkage: ECHILD includes the ILR, but young people who were captured in ILR would not have been linked to HES unless they also had an NPD record. However, within the school inception cohort (cohort 4, i.e. pupils attending school in 2001/02, who all should have been eligible for linkage), 11.8% of pupils in NPD did not have a linked HES record. Some of these pupils will have had a record in PDS but not in HES: overall, 2% of individuals in ECHILD with a TPI in PDS had no HES record). However, the vast majority of unlinked pupils in this cohort would have been excluded in ECHILD due to National Data Opt Outs. Published statistics show that Opt Out rates vary according to age, region and sex: as of 2023, when Opt Outs were applied to ECHILD, 4.4% of the population aged under 40 had opted out (5.0% of females and 3.8% of males).[26] Opt out rates increase by age, ranging from 1.3% of those aged 0-9 to 6.9% of those aged 30-39, and are more common in those living in the least deprived areas, and amongst those living in London. NHS England do not publish statistics on Opt Out rates by ethnicity; ECHILD is the first study to highlight substantial variation in Opt Out rates by ethnic group.[27]

In our attempt to estimate the rate of false matches within ECHILD, we found that a small proportion (<0.2%) of children who had died had a subsequent education record, which we assume indicates a false match. These records were likely to have missing data on other key variables, suggesting that poor data quality was driving the linkage error. We also found that 1% of patients in PDS linked to more than one pupil. This could be due to different children sharing the same set of identifiers (i.e. where the linkage algorithm could not distinguish between two children with the same combination of names, date of birth, sex and postcode), which would have resulted in at least one false match. Here, we are assuming that the internal linkage (or deduplication) within both PDS/HES and the NPD is accurate, i.e. we rely on the approaches that are used to assign unique identifiers to track the same individual through the dataset.[25] Previous analyses of the effect of updating the deduplication algorithm used for HES have described how this can lead to changes in entities, where records for the same individual are ‘split’, or records for different individuals are ‘merged’.[28] This phenomenon is not random, affecting specific subgroups more than others.[29] No published description or evaluation of NPD internal linkage algorithm is currently available.

The issue of multiple unique identifiers for the same individual within HES and the knock-on effects for linkage across different datasets is exemplified by the lower linkage rates between HES and NPD for those born in 1997-2002. These are likely explained by the fact that birth records were captured in HES for the first time in 1997 (when data collection began). Between 1997 and 2002, birth records and subsequent health records for the same child were more likely to have been assigned two different TPIs (babies were not assigned NHS numbers at birth until October 2002), resulting in the same patient being represented by more than one TPI (at least one of which would therefore not have been linked to an aPMR, since the linkage algorithm required that different TPIs could not be linked to the same aPMR). This ‘splitting’ of patients into multiple unique records also explains the unrealistically high numbers of births during this period, and has previously been recognised as a barrier to creating birth cohorts in HES prior to 2003.[30]

‘Population level’ datasets such as ECHILD often under-represent specific groups. Irrespective of the mechanisms by which individuals are excluded (e.g. Opt outs, linkage errors or not interacting with services), those in minoritised ethnic groups and those living in more deprived areas are most affected. This differential linkage has long been observed in the literature, including in an evaluation of a previous linkage algorithm for NPD and HES, and between mothers and babies in ECHILD.[8, 10, 31] Differential linkage has clear implications for inclusivity and representativeness of population-level datasets created by linking administrative data. Linkage evaluations such as the one we describe here are therefore critical for understanding who is and who is not included in analyses of these data.

There are a number of potential solutions to these challenges.

### Recommendations for data providers

First, maintaining public trust in the use of pseudonymised data for research is critical to avoiding the biases introduced by National Data Opt Outs, and better data on who chooses to opt out is required for researchers to quantify or mitigate the associated reduction in research quality.[32] A 2025 public consultation on data conducted by NHS England and the Department of Health and Social Care identified that members of the public want an opt out system that balances the need for choice with the need for data to be used for the greater good. However, it is unclear how many people opt back in to the use of their data once they have opted out.[32]

Second, data linkers can supply information about the linkage quality as part of routine data provision and enrichment.[33] Such reporting should provide clear eligibility denominators, and indicators of missed matches and potential false matches, ideally stratified by key characteristics (e.g., by deprivation, geography, and ethnicity). Record-level metadata (e.g., linkage route/strength, ambiguity flags) can support sensitivity analyses and transparent interpretation of subgroup estimates. More broadly, linkage quality should be communicated as part of a data-to-evidence pathway with clear, usable metadata and a feedback loop to data owners, so that analysts can anticipate where bias is likely and select proportionate mitigation strategies, supporting evidence that is fit to inform action towards healthier, fairer outcomes for all. Encouragingly, ongoing methodological improvements in NHS England’s person-level deduplication/linkage infrastructure illustrate that linkage quality is not static and can improve over time. For example, the total number of registered patients in England in PDS changed from 63.8 million in March 2025 to 63.7 million in March 2026; this difference is driven by ongoing work to address patient list inflation. However, these advances will only translate into better evidence if their implications for linkage error and representativeness are routinely quantified and communicated to data users.

### Recommendations for researchers

Researchers can treat linkage as a measurable source of selection and measurement error, making it visible and analysable in downstream research. While end users of linked datasets often have little influence over linkage processes or accuracy, or who is included in population-level data sources, it is important that researchers consider the implications for their analyses. As a minimum, researchers should carefully consider the selection of the denominator used in their study and the likely impact of biases arising from the inclusion or exclusion of unlinked records. For example, studies using ECHILD to examine health outcomes in children at school may restrict their school cohorts to pupils who appear at least once within linked health data, to avoid the inclusion of pupils in the denominator who could never have a record of the outcome (e.g. mental health services referrals, ACE attendances, or admissions).[34] By reporting rates of non-linkage (excluded pupils) by gender and ethnicity, researchers have demonstrated that inclusion of these pupils in the denominator would have downwardly biased outcomes for groups with lower linkage rates (e.g. Black Caribbean pupils) relative to those with higher linkage rates.[27] However, exclusion of unlinked pupils will have implications for the representativeness of the cohort. Other approaches, such as multiple imputation, could be considered when linkage supports capture of covariate data.

### Limitations

Our study has several limitations. First, although National Data Opt Outs are likely to explain a substantial proportion of non-linkage between education and health records, we were unable to directly quantify their contribution because demographic information on opted-out individuals was not provided by NHS England. Second, the observed differences by ethnicity may reflect a combination of factors, including differential data quality, migration, service use, private education, and opt-out behaviour; the relative contribution of these mechanisms could not be disentangled within this study. Finally, our assessment of false matches using children who died before school age provides only a lower-bound estimate of linkage error, as it identifies one specific type of implausible linkage and does not capture all potential false matches within the dataset.

## Conclusion

‘Population level’ datasets, including ECHILD, often under-represent specific groups. Irrespective of the mechanisms by which individuals are excluded (e.g. Opt outs, linkage errors or not interacting with services), those in minoritised ethnic groups and those living in more deprived areas are most affected. Our findings demonstrate that evaluations of linked administrative data should consider both linkage quality and population representation. For many research applications, understanding who is excluded from the linked resource may be as important as understanding the technical performance of the linkage algorithm itself.

## Acknowledgements

ECHILD uses data from the Department for Education (DfE). The DfE does not accept responsibility for any inferences or conclusions derived by the authors. This work uses data provided by patients and collected by the National Health Service as part of their care and support. Source data can also be accessed by researchers by applying directly to NHS England.

We are grateful to the Office for National Statistics (ONS) for providing the trusted research environment for ECHILD. ONS agrees that the figures and descriptions of results in the attached document may be published. This does not imply ONS’ acceptance of the validity of the methods used to obtain these figures, or of any analysis of the results.

The DfE, NHSE and ONS do not accept responsibility for any inferences or conclusions derived by the authors.

We thank all the children, young people, parents and carers who contributed to ECHILD. We gratefully acknowledge all children and families whose de-identified data are used in this research. We would also like to acknowledge the contribution of the wider ECHILD support and programme management.

## Ethics

Ethical approval was granted by the National Research Ethics Service (17/LO/1494) and NHS Health Research Authority Research Ethics Committee (20/EE/0180 and 21/SW/0159).

## Data Availability Statement

Data in ECHILD are available on request through www.echild.ac.uk for access within the Office for National Statistics Secure Research Service for approved researchers. Data cannot be made publicly available due to disclosure control.

## AI Disclosure Statement

The authors used Claude AI to proofread the manuscript prior to submission. The output was reviewed and verified by the authors.

## Supplementary material

Appendix 1: Component datasets within ECHILD and associated time periods

Appendix 2: Linkage process

Appendix 3: Unique pupils, patients and links within ECHILD

Appendix 4: Flow diagram for Cohort 1: HES to NPD linkage: Individuals born between September 1984 and August 2017.

Appendix 5: Characteristics of unlinked HES records from Cohort 1 and Cohort 2

Appendix 6: Flow diagram for Cohort 2: HES to NPD linkage: Birth Cohort April 2003 to August 2017

Appendix 7: Flow diagram for Cohort 3: NPD to HES linkage: Any pupils captured in NPD

Appendix 8: Characteristics of unlinked NPD records from Cohort 3 and Cohort 4

Appendix 9: Flow diagram for Cohort 4: NPD to HES linkage: School inception 2001/02

