## Appendices 1-9 for "Evaluation of linkage between health, education and social care administrative data for 21 million children in England"

**Appendix 1: Component datasets within ECHILD and associated time periods**

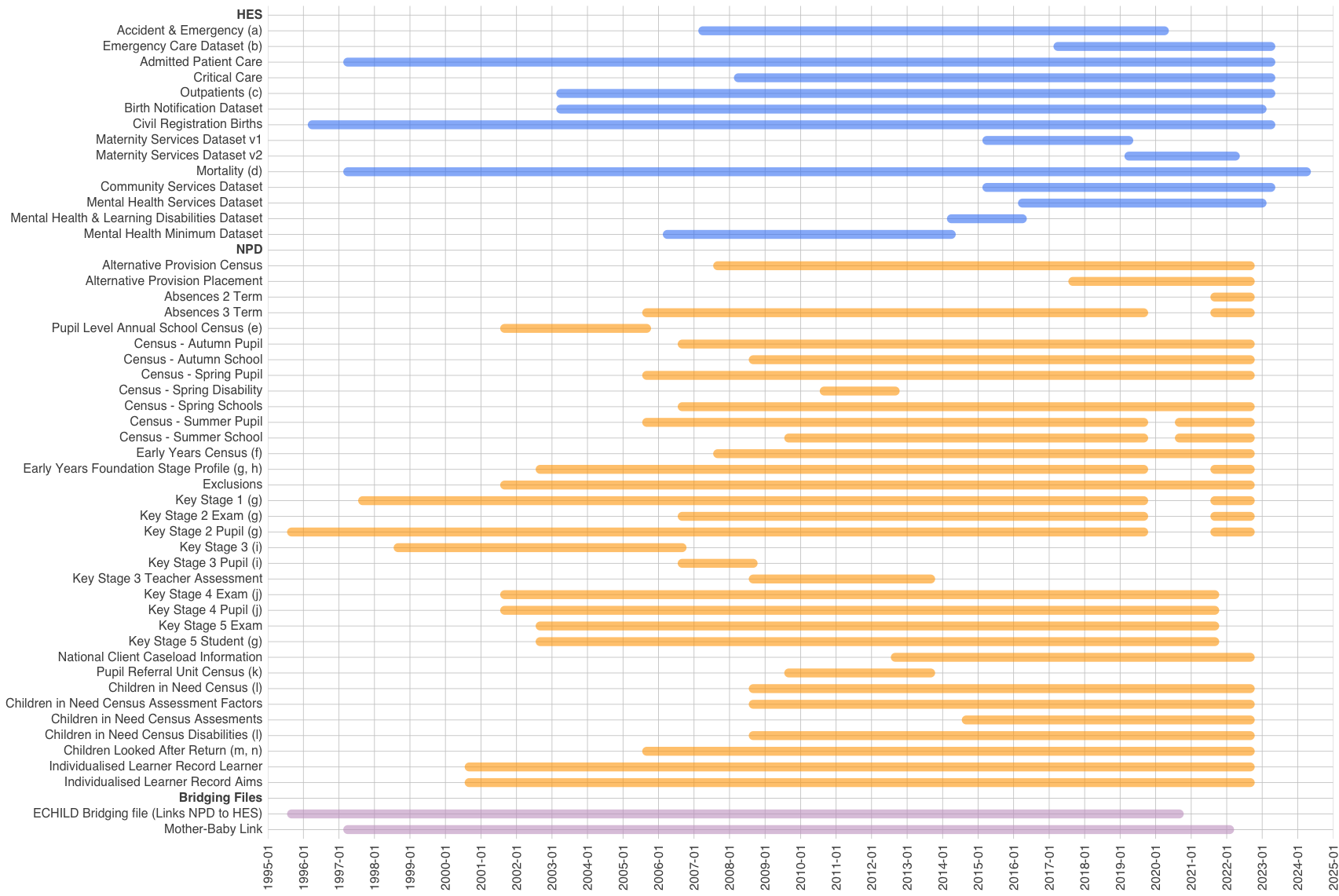

**Reference table for Appendix 1**

| Key | Detail |
| --- | --- |
| a | Data from 2007/08 to 2011/12 were classified as “experimental” by NHS England. |
| b | ECDS data from October 2017 to 2019/20 are considered pilot data, with ECDS formally replacing HES A&E in 2020/21. |
| c | Data from 2003/04 to 2007/08 were classified as “experimental” by NHS England. |
| d | ONS Mortality data were first linked to HES in January 1998. |
| e | From 2001/02 to 2004/05 annual census exists, from 2005/06 onwards census data are termly. |
| f | The Early Years Census included 3- to 4-year-olds between 2007/08 and 2012/13. From 2013/14 it also includes 2- to 4-year-olds. |
| g | Not collected in 2019/2020 and 2020/21 academic years due to COVID. |
| h | Partial coverage as between the 2002/03 and 2005/06 academic years, data only on a 10% sample of children. |
| i | Key Stage 3 assessments ceased after 2012/13. |
| j | Institutional identifiers not included in 2019/20 and 2020/21 academic years due to restrictions on comparing institutional performance during the pandemic years. |
| k | PRU Census data were subsumed into the School Census Pupil Level table from 2013/14. |
| l | CIN: Linkage to NPD is incomplete prior to September 2012. |
| m | Partial coverage of population as between 01/04/1992 & 31/03/2003, CLA data were only collected for a one-third sample (i.e., children with a day of birth divisible by 3). |
| n | CLA: Linkage to NPD from 1 April 2005 onwards. |

**Appendix 2: Linkage process**

### Data sources

#### The Department for Education’s National Pupil Database (NPD)

Within NPD, each record is associated with identifiers (forename, surname, gender, date of birth, postcode) relating to the pupil’s details as known by the submitting organisation at the time the data were submitted. When a pupil first attends a state-funded school in England e.g., nursery or primary school, or has an education, health and care plan (EHCP) put in place, they are allocated a ‘Unique Pupil Number’ (UPN), which remains with the pupil throughout their school career regardless of any change in school or local authority. Social care data is included in the NPD for children who have a UPN. Children receiving social care preschool entry who never have social care during their school years are therefore not included in ECHILD. UPNs facilitate the transfer of school-based education and attainment data between schools, local authorities and central government and are stored within the NPD.

DfE additionally assigns each record in NPD an anonymised Pupil Matching Reference (aPMR). An aPMR is an identifier which is not in itself meaningful: it does not reveal the identity of the pupil. aPMRs are assigned to all records in NPD such that one aPMR should represent one pupil and each pupil should have only one aPMR. This allows records for each pupil to be identified across NPD, both between different datasets and over time, without revealing their identity. As a result, NPD contains a longitudinal record of pupils’ names, addresses, and (potentially) genders over time.

#### NHS England’s Personal Demographics Service and Master Person Service

NHS England operates the Personal Demographics Service (PDS), a national electronic database of demographic data for patients accessing care in England or services funded by the NHS in England. PDS contains identifiers (forename, surname, gender, date of birth, postcode) and NHS number. Since October 2002, all babies born in England (and Wales) have been assigned a distinct NHS number. Prior to this, babies were not given an NHS number until they were officially registered at the Registrar of Births and Deaths (which could take six weeks). NHS numbers are otherwise assigned at the first time of accessing NHS services. PDS holds a longitudinal record of name and address changes made to NHS services in England over time. This means there may be many records for each person in PDS but all records for a person should be assigned the same NHS number (with some exceptions, e.g. adoptions and gender reassignment).

NHS England also manage the Master Person Service (MPS), which takes a record of identifiers and attempts to link this to a PDS record, allowing for some errors and missingness in the recording of identifiers. If this fails and the identifiers are sufficiently complete, MPS attempts to link records against a secondary store (MPS bucket) of natural identifiers of persons who previously had contact with the NHS in England and do not have an NHS number. These persons are assigned an “MPS ID”.

The MPS returns a “Person ID” using either:

1.	NHS number, if a valid link is found in PDS; or,

2.	MPS ID if no link is found in PDS but a valid link is found in the MPS bucket; or,

3.	No value if no link is found in either PDS or the MPS bucket.

The Person ID is then encrypted to generate a Token Person ID (TPI), which is not meaningful and does not reveal the person’s identity. Within ECHILD, the TPI can be used to follow records belonging to the same patient.

### Linking DfE NPD aPMRs to NHS England TPIs

For the purposes of the ECHILD linkage, the following simplifying assumptions were made:

1. Each aPMR represents at most one real person within NPD;
2. Each TPI represents precisely one real person within NHS England data collections;
3. Each real person represented within NHS England data collections has precisely one TPI.

Essentially, whilst we assume TPIs are perfectly allocated, we only require that aPMRs are not shared. That is, the same real person is permitted to have more than one aPMR.

This linkage task resulted in “N-to-one” links between aPMRs and TPIs: each aPMR linked to at most one TPI but a TPI may have linked to more than one aPMR.

#### Linkage Stage 1: Exact link

An initial, simple, linkage stage was used to avoid over-burdening the more resource-intensive MPS trace. Each valid record (e.g., no blank entries) in the NPD linkage dataset was compared to all records in a prepared extract from PDS. A sequential, two part, approach was used. Only records belonging to aPMRs not linked after Part A progressed to Part B. In both parts, an aPMR was considered linked only if all of its associated linkage records were linked to at most one TPI and at least one record was linked to a TPI.

Part A: Full forename

A record was deemed linked if compared records had equal (non-empty) values across all of full forename, full surname, full date of birth, full postcode and gender.

Part B: Partial forename

A record was deemed linked if compared records had equal (non-empty) values across all of first four characters of forename, full surname, full date of birth, full postcode, and gender.

#### Linkage Stage 2: MPS Trace

Only records belonging to aPMRs not linked after Linkage Stage 1 were submitted to MPS, aiming to return a PersonID as described above.

### Linkage results

DfE provided 430M sets of identifiers to NHS England, covering 22.8M aPMRs (this figure differs from the 25.3M aPMRs in ECHILD, as identifiers were only extracted for linkage from a selected set of tables). NHS England linked 22.6M (99.1%) aPMRs to a TPI, in the following stages:

- Stage 1 (Exact link) Part A: Full forename. 20.1M (88.2%) aPMRs linked
- Stage 1 (Exact link) Part B: Partial forename. 1.3M (5.7%) aPMRs linked
- Stage 2 (MPS trace): 1.2M (5.3%) aPMRs

Less than 200,000 aPMRs remained unlinked at the end of Stage 2: ~160,000 were not resolvable to any TPI; ~30,000 resolved to more than one TPI and (due to the simplifying assumptions) these were considered invalid and so no link was recorded for any of these aPMRs.

**Appendix 3: Unique pupils, patients and links within ECHILD**

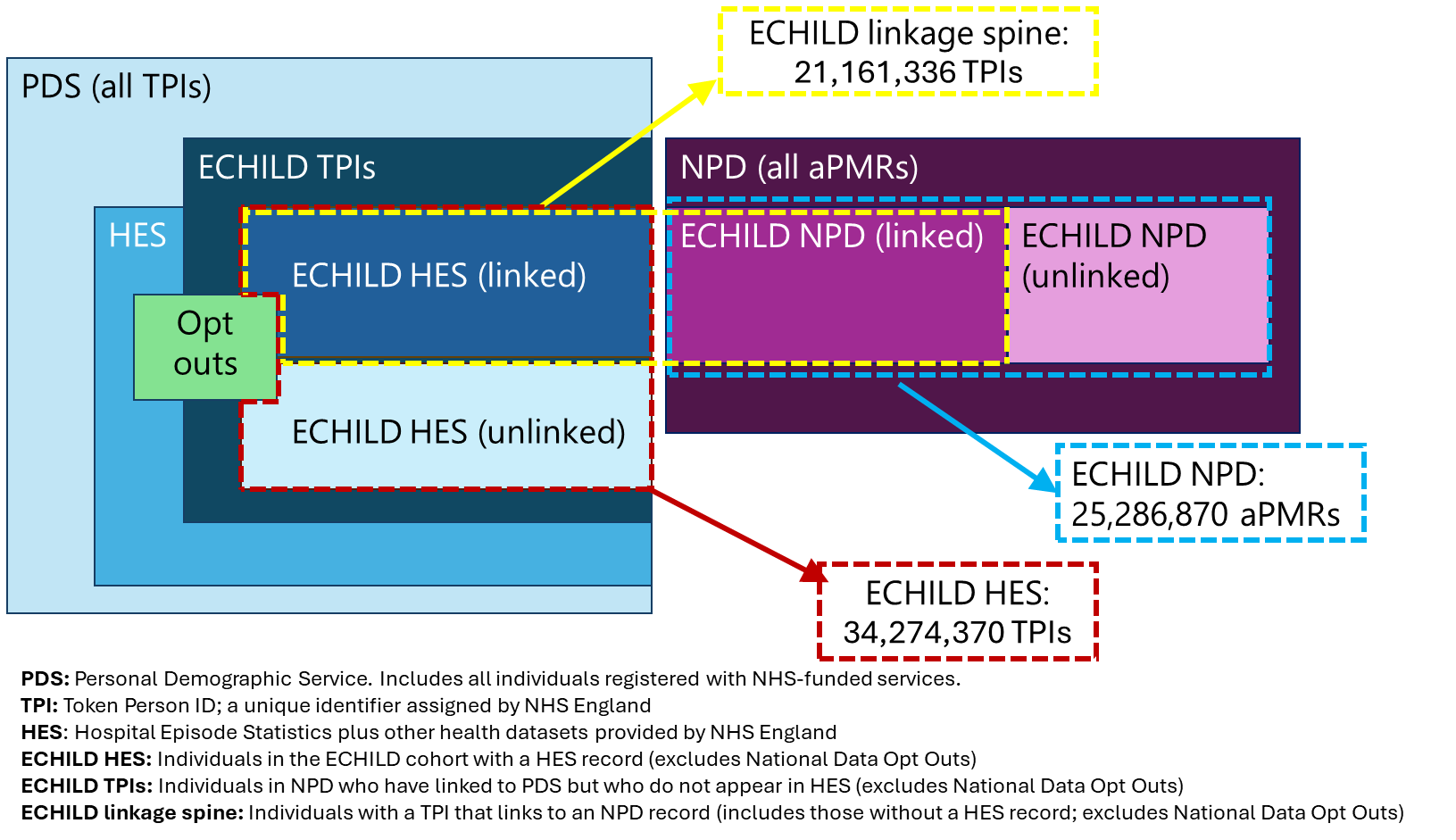

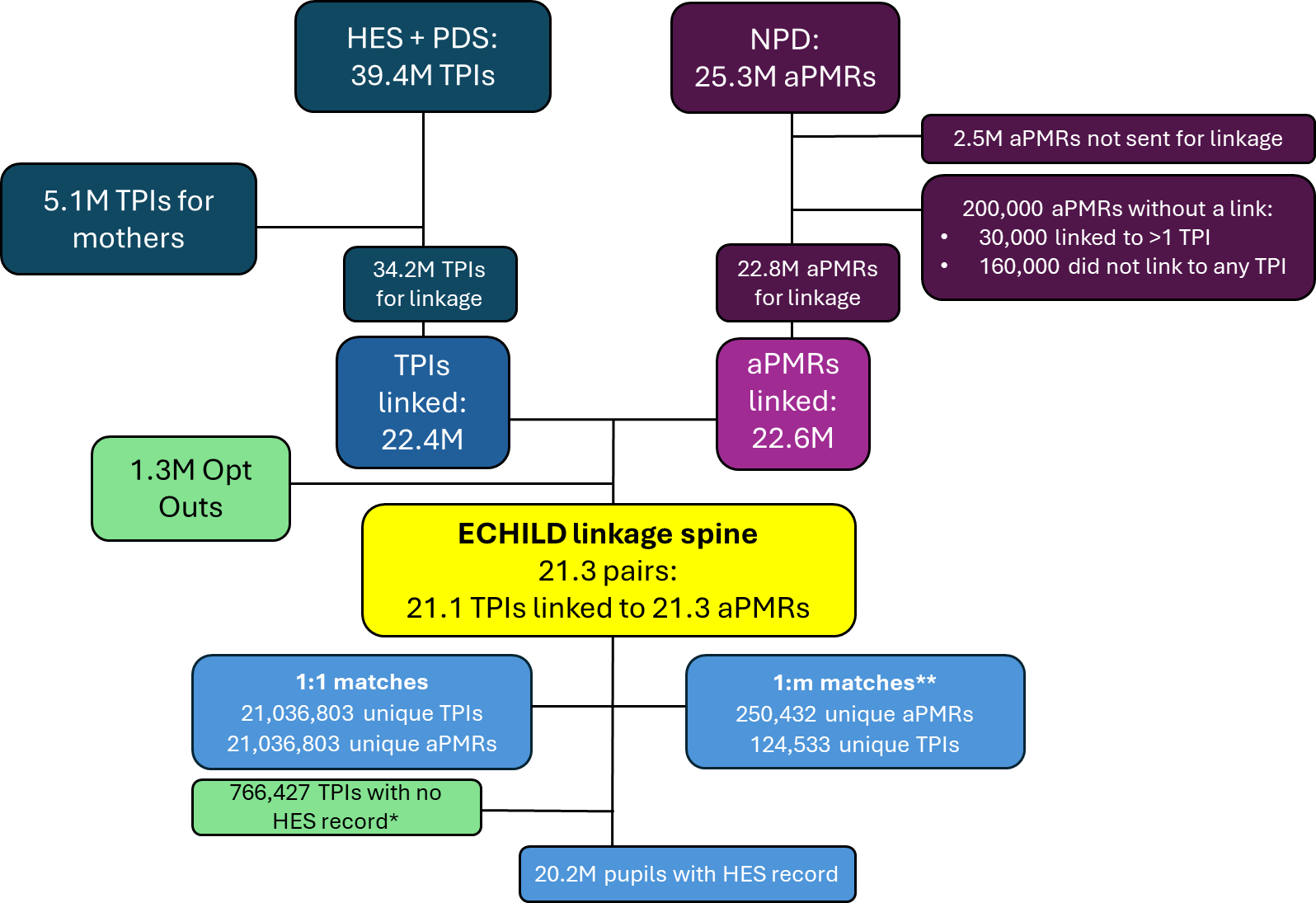

**PDS:** Personal Demographic Service. Includes all individuals registered with NHS-funded services (includes those with birth notifications).

**TPI:** Token Person ID; a unique identifier assigned by NHS England

**HES:** Hospital Episode Statistics plus other health datasets provided by NHS England

**ECHILD HES:** Individuals in the ECHILD cohort with a HES record (excludes National Data Opt Outs)

**ECHILD TPIs:** Individuals in NPD who have linked to PDS but who do not appear in HES (excludes National Data Opt Outs)

**ECHILD linkage spine:** Individuals with a TPI that links to an NPD record (includes those without a HES record; excludes National Data Opt Outs)

***** Some patients were included in PDS but had no HES record

** TPIs were assumed to be unique but aPMRs were not: each aPMR was therefore linked to at most one TPI, but a TPI may have linked to more than one aPMR

**Appendix 4: Flow diagram for Cohort 1: HES to NPD linkage: Individuals born between September 1984 and August 2017.**

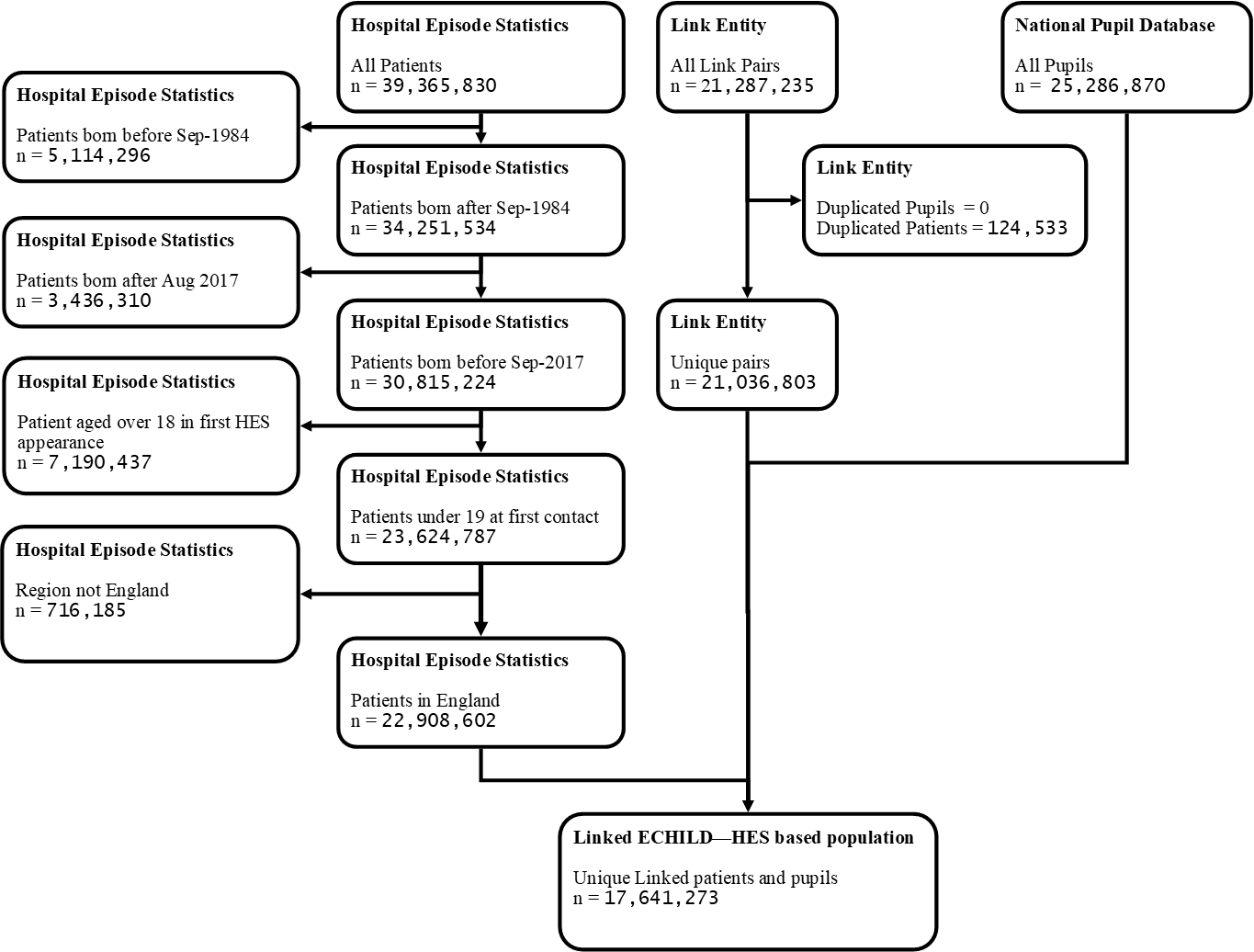

**Appendix 5: Characteristics of unlinked HES records from Cohort 1 and Cohort 2**

|  |  | **Cohort 1: Full HES cohort N=22,908,602** | | **Cohort 2: Birth cohort (2003-2017) N=8,729,009** | |
| --- | --- | --- | --- | --- | --- |
|  |  | N unlinked records | % Unlinked records | N unlinked records | % Unlinked records |
| **Total** |  | 5,267,329 | 23.0% | 814,577 | 9.3% |
| **Sex** | Male | 2,691,526 | 22.8% | 330,093 | 8.9% |
|  | Female | 2,465,919 | 22.5% | 307,996 | 8.8% |
|  | Missing | 101,624 | 94.9% | 176,488 | 11.6% |
|  | Undetermined | 8260 | 90.7% | - | - |
| **Ethnicity** | British (White) | 1,037,397 | 9.0% | 275,449 | 6.6% |
|  | Irish (White) | 18,453 | 32.3% | 4292 | 19.4% |
|  | White* (prior to 2000/01) | 691,105 | 60.8% | - | - |
|  | Any other White background | 276,405 | 23.9% | 95,169 | 17.7% |
|  | African (Black or Black British) | 92,854 | 19.6% | 27,979 | 12.4% |
|  | Caribbean (Black or Black British) | 38,587 | 21.8% | 6042 | 9.6% |
|  | Any other Black background | 41,561 | 21.7% | 5577 | 11.7% |
|  | White and Asian (Mixed) | 13,506 | 11.9% | 5319 | 8.6% |
|  | White and Black African (Mixed) | 9082 | 12.6% | 3437 | 9.0% |
|  | White and Black Caribbean (Mixed) | 12,362 | 9.2% | 4161 | 6.7% |
|  | Any other Mixed background | 35,496 | 15.2% | 12,459 | 10.8% |
|  | Bangladeshi (Asian or Asian British) | 26,007 | 13.0% | 5739 | 6.1% |
|  | Indian (Asian or Asian British) | 73,429 | 16.3% | 19,583 | 10.1% |
|  | Pakistani (Asian or Asian British) | 83,478 | 13.9% | 19,093 | 7.3% |
|  | Any other Asian background | 61,237 | 18.0% | 14,032 | 11.7% |
|  | Chinese (other ethnic group) | 25,476 | 29.4% | 5727 | 16.7% |
|  | Any other ethnic group | 196,907 | 30.4% | 24,264 | 14.8% |
|  | Not stated / Not known | 91,868 | 16.1% | 82,188 | 11.3% |
|  | Missing | 2,442,119 | 51.9% | 204,067 | 11.4% |
| **Region** | London | 962,968 | 26.5% | 76,139 | 12.0% |
|  | East Midlands | 195,518 | 12.0% | 14,940 | 5.4% |
|  | East of England | 312,527 | 14.7% | 23,795 | 7.5% |
|  | Merseyside (until 1998/99) | 27,020 | 76.1% | - | - |
|  | North East | 143,792 | 14.4% | 9725 | 4.6% |
|  | North West | 421,651 | 15.2% | 50,410 | 8.9% |
|  | South East | 559,213 | 17.5% | 44,831 | 7.3% |
|  | South West | 307,429 | 16.2% | 21,575 | 5.2% |
|  | West Midlands | 392,935 | 17.4% | 24,289 | 6.5% |
|  | Yorkshire and Humber | 256,831 | 12.7% | 19,471 | 5.2% |
|  | No fixed abode | 34,370 | 87.9% | 399 | 36.9% |
|  | Unknown | 1,653,075 | 72.1% | 529,003 | 10.7% |
| **IMD decile** | Least deprived 10% | 296,919 | 15.5% | 10,994 | 5.7% |
|  | Less deprived 10-20% | 289,783 | 15.3% | 10,603 | 5.4% |
|  | Less deprived 20-30% | 292,055 | 15.9% | 11,043 | 5.7% |
|  | Less deprived 30-40% | 294,548 | 16.1% | 11,937 | 6.0% |
|  | Less deprived 40-50% | 312,878 | 16.6% | 13,625 | 6.4% |
|  | More deprived 10-20% | 482,448 | 17.7% | 23,155 | 7.4% |
|  | More deprived 20-30% | 410,059 | 17.8% | 20,160 | 7.2% |
|  | More deprived 30-40% | 350,661 | 17.2% | 17,285 | 6.9% |
|  | More deprived 40-50% | 311,081 | 17.2% | 15,381 | 6.5% |
|  | Most deprived 10% | 523,995 | 18.8% | 26,121 | 7.1% |
|  | Unknown | 1,702,902 | 90.4% | 654,273 | 10.4% |
| **Gestational age at birth (weeks)** | <24 |  |  | 4803 | 59.6% |
|  | 24-31 |  |  | 27,502 | 18.2% |
|  | 32-33 |  |  | 7282 | 12.9% |
|  | 34-36 |  |  | 30,852 | 10.1% |
|  | 37-38 |  |  | 95,770 | 8.3% |
|  | 39 |  |  | 105,682 | 7.9% |
|  | 40 |  |  | 128,696 | 7.8% |
|  | 41-43 |  |  | 104,418 | 7.7% |
|  | 44+ |  |  | 802 | 7.9% |
|  | Missing |  |  | 316,052 | 11.4% |
| **Birthweight** | < 500g |  |  | 5359 | 48.9% |
|  | 500-1499g |  |  | 21,208 | 30.5% |
|  | 1500-2499g |  |  | 40,919 | 10.7% |
|  | 2500-3499g |  |  | 269,417 | 8.2% |
|  | 3500g+ |  |  | 186,685 | 7.4% |
|  | Missing |  |  | 290,989 | 11.8% |
| **Maternal age (years)** | <20 |  |  | 29,183 | 9.6% |
|  | 20-29 |  |  | 217,973 | 8.5% |
|  | 30-39 |  |  | 231,855 | 9.1% |
|  | 40+ |  |  | 19,746 | 9.2% |
|  | Missing |  |  | 315,820 | 10.2% |

* Prior to 2000/01, only “White” was available; subsequently, White British, Irish or Any other background became available

**Appendix 6: Flow diagram for Cohort 2: HES to NPD linkage: Birth Cohort April 2003 to August 2017**

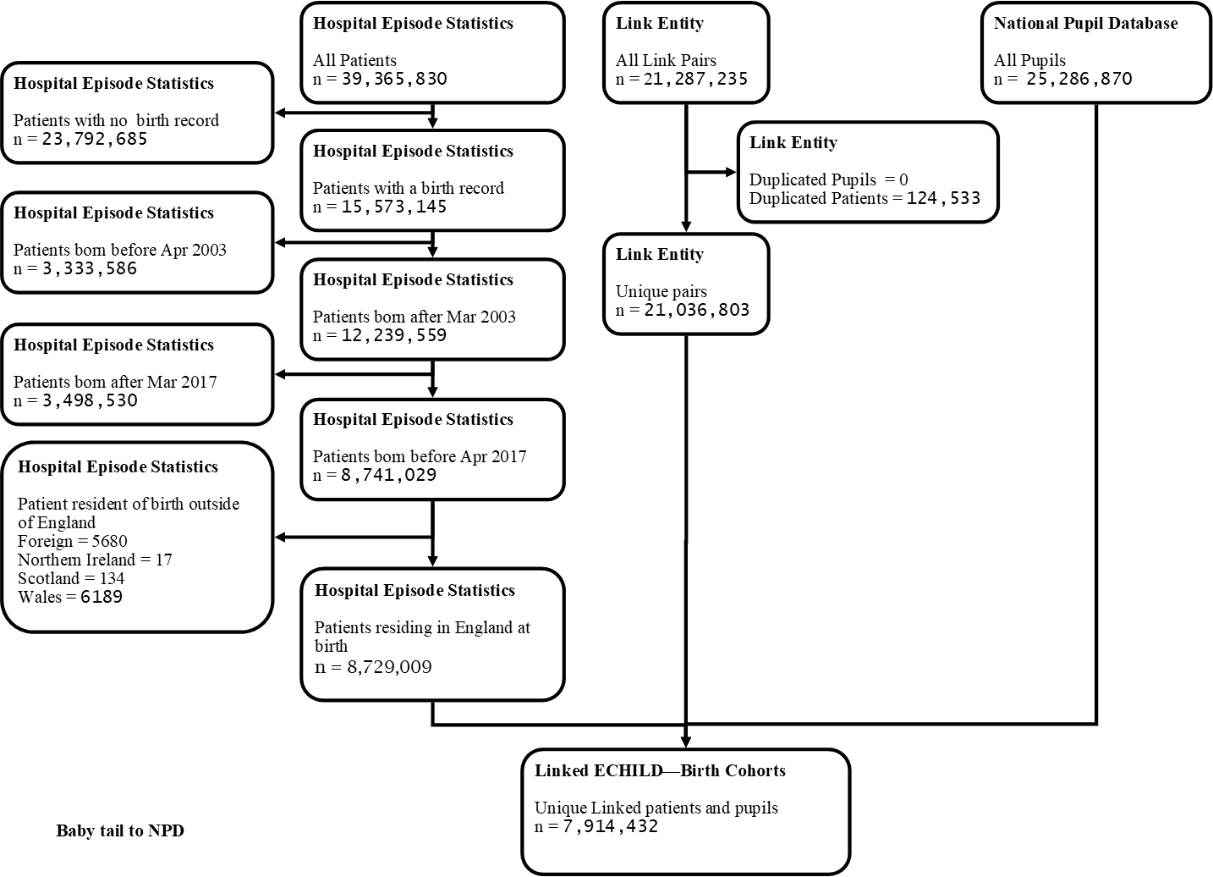

**Appendix 7: Flow diagram for Cohort 3: NPD to HES linkage: Any pupils captured in NPD**

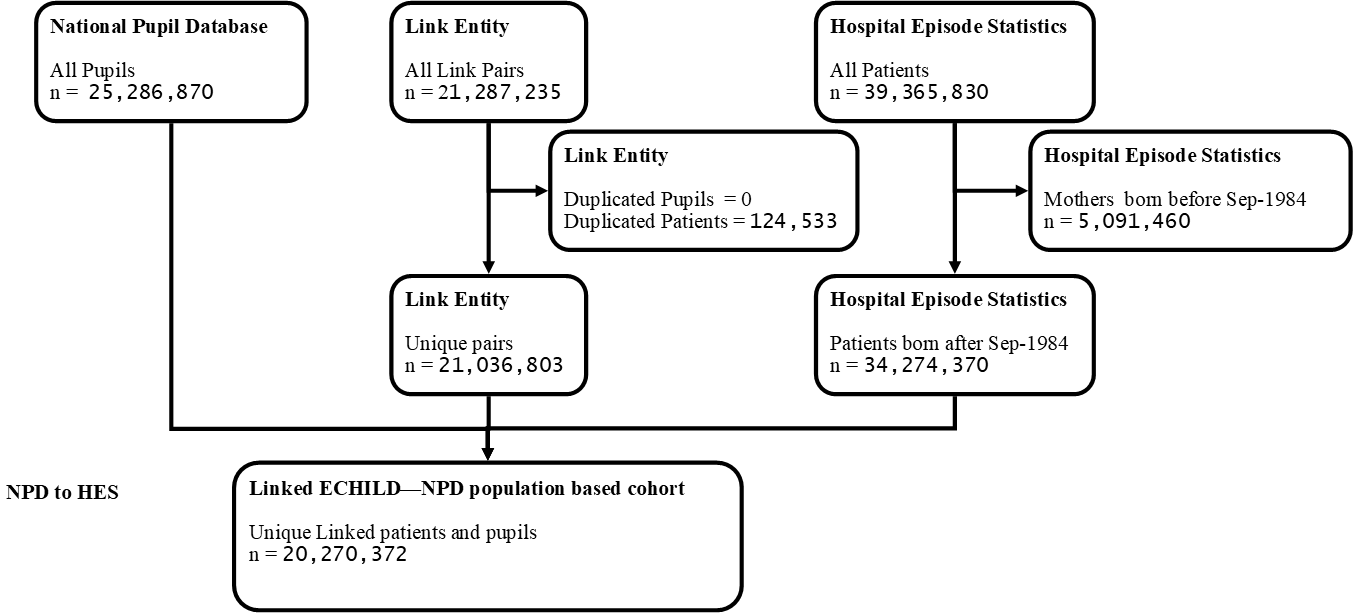

**Appendix 8: Characteristics of unlinked NPD records from Cohort 3 and Cohort 4**

|  |  | **Cohort 3: Full NPD cohort** | | **Cohort 4: School inception cohort** | |
| --- | --- | --- | --- | --- | --- |
|  |  | N unlinked records | % Unlinked records | N unlinked records | % Unlinked records |
| **Total** |  | 5,016,498 | 19.8% | 895,262 | 11.8% |
| **Gender** | Male | 1,909,969 | 15.5% | 435,320 | 11.3% |
|  | Female | 1,904,191 | 16.2% | 459,942 | 12.4% |
|  | Unknown | 1,202,338 | 99.6% |  |  |
| **Ethnicity** | White | 1,154,055 | 8.0% | 698,862 | 10.6% |
|  | Black | 194,572 | 19.0% | 70,227 | 23.8% |
|  | Asian | 282,691 | 14.6% | 81,002 | 17.4% |
|  | Mixed | 115,755 | 12.0% | 870 | 17.4% |
|  | Chinese | 28,342 | 27.7% | 5395 | 21.4% |
|  | Any other ethnic group | 86,626 | 22.6% | 35,343 | 19.5% |
|  | Missing | 3,154,457 |  | 3563 | 10.5% |
| **IDACI decile** | Most deprived | 350,229 | 12.0% | 125,676 | 12.8% |
|  | 2 | 286,177 | 11.3% | 103,051 | 11.8% |
|  | 3 | 239,044 | 10.6% | 91,231 | 11.4% |
|  | 4 | 213,667 | 10.2% | 83,754 | 11.2% |
|  | 5 | 196,639 | 9.9% | 80,338 | 11.4% |
|  | 6 | 182,699 | 9.7% | 78,834 | 11.4% |
|  | 7 | 177,749 | 9.4% | 77,380 | 11.3% |
|  | 8 | 173,879 | 9.4% | 78,653 | 11.6% |
|  | 9 | 174,681 | 9.5% | 79,666 | 11.9% |
|  | Least deprived | 186,227 | 10.3% | 84,197 | 12.8% |
|  | Unknown | 2,835,507 | 67.0% | 12,482 | 13.3% |
| **Region** | London | 792,359 | 21.3% | 178,629 | 17.2% |
|  | East Midlands | 280,493 | 13.8% | 71,341 | 10.7% |
|  | East of England | 405,238 | 15.2% | 101,921 | 12.4% |
|  | North East | 129,843 | 11.6% | 37,344 | 9.0% |
|  | North West | 465,741 | 14.3% | 127,231 | 11.5% |
|  | South East | 652,937 | 16.6% | 131,607 | 11.5% |
|  | South West | 311,755 | 13.9% | 64,112 | 10.4% |
|  | West Midlands | 382,297 | 14.5% | 92,352 | 10.6% |
|  | Yorkshire | 302,086 | 12.7% | 79,834 | 9.8% |
|  | Unknown | 1,293,749 | 98.1% | 10,891 | 11.6% |
| **Free School Meals** | No record | 4,475,356 | 23.2% | 749,451 | 11.9% |
|  | At least one record | 541,142 | 9.0% | 145,811 | 11.6% |
| **Special Educational Needs and Disability** | No record | 4,456,403 | 24.3% | 743,142 | 12.4% |
|  | At least one record | 560,095 | 8.0% | 152,120 | 9.6% |
| **National Curriculum Year group** | Reception |  |  | 30,861 | 10.2% |
|  | 1 |  |  | 63,807 | 11.2% |
|  | 2 |  |  | 68,089 | 11.8% |
|  | 3 |  |  | 68,758 | 11.8% |
|  | 4 |  |  | 69,567 | 11.7% |
|  | 5 |  |  | 69,463 | 11.7% |
|  | 6 |  |  | 70,871 | 11.5% |
|  | 7 |  |  | 70,094 | 11.4% |
|  | 8 |  |  | 69,864 | 11.4% |
|  | 9 |  |  | 69,632 | 11.6% |
|  | 10 |  |  | 72,090 | 11.9% |
|  | 11 |  |  | 73,299 | 12.5% |
|  | 12 |  |  | 71,617 | 13.1% |
|  | 13 |  |  | 26,676 | 15.3% |
|  | Missing |  |  | 574 | 15.8% |
| **Mother tongue** | English |  |  | 752,944 | 11.1% |
|  | Other than English |  |  | 137,473 | 18.9% |
|  | Not known; believed to be English |  |  | 3802 | 11.2% |
|  | Not known; believed to be other |  |  | 755 | 19.5% |
|  | Not recorded |  |  | 288 | 15.2% |

IDACI = Income Deprivation Affecting Children Index

**Appendix 9: Flow diagram for Cohort 4: NPD to HES linkage: School inception 2001/02**

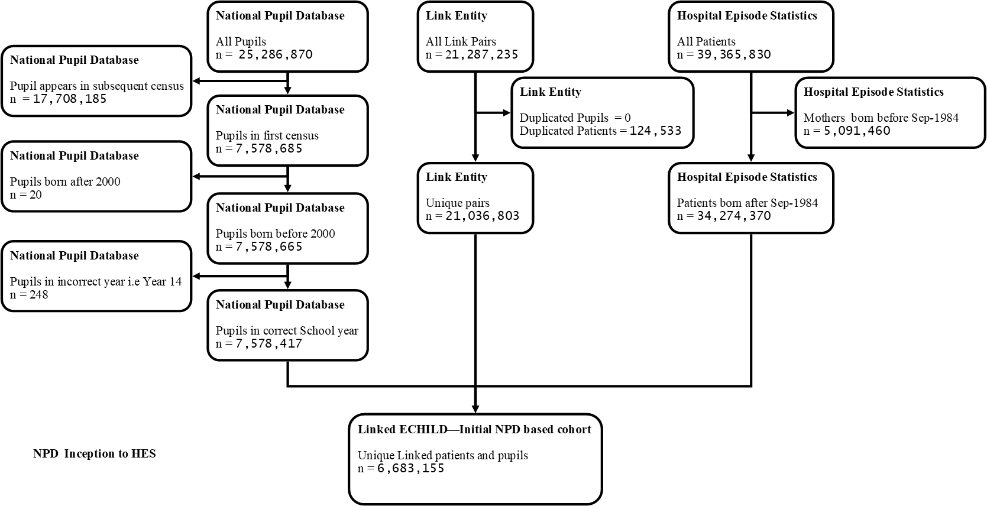
